# Isolating the genetic component of mania in bipolar disorder

**DOI:** 10.1101/2024.08.30.24312859

**Authors:** Giuseppe Pierpaolo Merola, Johan Zvrskovec, Rujia Wang, Yuen Kaye Li, Giovanni Castellini, Valdo Ricca, Jonathan Coleman, Evangelos Vassos, Gerome Breen

## Abstract

**Objective:** Bipolar disorder typically features episodes of mania and depression, frequently accompanied by psychosis. While progress has been made in understanding the genetics of depression and psychosis, mania remains underexplored.

**Methods:** We employed Genomic Structural Equation Modeling to subtract the genetic effects of schizophrenia and major depressive disorder (MDD) from bipolar disorder to identify a genetic trait specific to mania.

**Results:** The SEM model revealed significant loadings for “mania” (0.67, p<0.001), “psychosis” (0.58, p<0.001), and “depression” (0.29, p<0.001) factors, with mania, MDD and schizophrenia explaining 45%, 8% and 34% of the variance in bipolar disorder, respectively. Seven significant genomic regions associated with mania were identified. Key regions include rs9834970 (3q12.1, previously associated with lithium response), rs6992333 (8q13.1, brain structure), and rs12206087 (6q14.3, intelligence and cortical surface). Additionally, mania exhibited distinct genetic correlations compared to bipolar disorder across psychiatric, substance abuse, somatic, social, and neurological traits, including significantly higher correlations with intelligence (r_g_=0.08 vs −0.07) and educational attainment (r_g_=0.17 vs 0.12), and an unexpected reduced correlation with risky sexual behavior (r_g_=0.14 vs 0.27).

**Conclusions:** These findings enhance understanding of bipolar disorder’s genetic architecture, potentially offering a more bipolar disorder-specific GWAS.

## Introduction

Mania is a cardinal symptom of bipolar disorder, a severe and relatively rare psychiatric condition affecting about 2% of the population, characterized by alternating phases of mania and depression (1). Mania involves sleeplessness, heightened mood and energy levels, irritability, increased psychomotor activity, rapid speech and thought, and, in extreme cases, delusions of grandeur and hallucinations (2). Hypomania, a milder form, has similar symptoms, without severe impairment or psychotic symptoms (3). Depressive episodes exhibit symptoms such as decreased energy, apathy, and anhedonia (2).

For individuals, mania can lead to impulsive spending, risky investments, and deteriorating relationships (4). Manic episodes also burdens families with emotional stress and caregiving challenges (5). Societally, the impact includes higher healthcare costs and the need for social support services (6). Acutely, second-generation antipsychotics are used to manage symptoms, and lithium and mood stabilizers provide maintenance therapy. However, treating mania presents significant challenges, both due to low compliance and clinical complexities. Antidepressants, often used when bipolar disorder is misdiagnosed as depression, can exacerbate mania (7). Additionally, existing treatments often have significant long-term side effects and/or partial effectiveness (8). Thus, there is a pressing need for better therapeutic and intervention options for mania in bipolar disorder, which could significantly improve patient outcomes (9).

Mood congruent and incongruent psychotic symptoms can appear during any phase of bipolar disorder (10). Thus, while bipolar disorder formally falls into the category of mood disorders under the DSM-5 and ICD-11 classifications (11,12), its symptomatology spans both the mood disorder and psychosis spectrum (13,14).

Bipolar disorder is multifactorial, with its etiology arising from both genetic and environmental or social factors (15) with a heritability between 60% and 85% (16). The most comprehensive genome-wide association study (GWAS) published to date (17) identified 64 single nucleotide polymorphisms (SNPs) and found strong genetic correlations with many psychiatric conditions, particularly schizophrenia (r_g_=0.68) and major depressive disorder (MDD; r_g_=0.44).

Despite extensive research into the genetics of bipolar disorder, specific attempts to study the genetics of its mania component have not yet been undertaken. This is a significant gap; as discussed earlier, mania is a cardinal and a disruptive symptom of bipolar disorder. Identifying genetic variants specifically associated with mania could provide crucial insights into its biological mechanisms, potentially leading to better discrimination of bipolar disorder among people with a first depressive episode (18).

This study aims to address this gap using a genomic structural equation modeling (GSEM) approach to perform a GWAS of a derived, latent factor specific to mania (19). This approach is based on the successful GWAS-by-subtraction on educational attainment to derive its non-cognitive component (20).

## Methods

### GWAS Summary Statistics and Quality Control

The analysis incorporated recent GWAS summary statistics for three major psychiatric disorders from large-scale studies of European ancestry samples. For MDD, data from Wray et al. (21), excluding 23andMe to ensure phenotypic accuracy (22), included 59,851 cases and 113,154 controls. Schizophrenia data from Trubetskoy et al. (23) included 74,776 cases and 101,023 controls. For bipolar disorder, data from Mullins et al. (17) comprised 41,917 cases and 371,549 controls. All summary statistics were accessed from the Psychiatric Genomics Consortium (PGC) website (24). Quality control and reference harmonization were performed to correct formatting issues and align alleles and effect directions. An INFO score threshold of 0.6 and a minor allele frequency (MAF) filter of 0.01 were applied to exclude low imputation quality associations and rare variants (25).

### Genomic Structural Equation Modeling

GSEM (19) was employed, which applies structural equation modeling (SEM) on GWAS summary statistics. SEM is a statistical technique that combines factor analysis and linear regression to examine relationships between observed variables and latent constructs. GSEM allows incorporation of genomic data into SEM. Our analysis was conducted using the R package “Genomic SEM” (19) in R 4.30 using the GWAS summary statistics for bipolar disorder, schizophrenia, and MDD. An SEM model (described below) was fitted to genomic covariance estimated by LD score regression (26) as implemented in the same R package, and using European ancestry LD scores based on the European ancestry subset of the 1000 Genomes Project sample (27), restricted to the HapMap3 variants only (28). Python 3.11.6 was employed for plotting.

### Double-GWAS-by-subtraction SEM Model

The structural equation model (Figure 1) was specified using the “lavaan” syntax in R (29). We defined three latent variables: the “subtracting” variables (*Depression* and *Psychosis*) and *Mania*. The primary focus was on *Mania* as the main latent factor, while the factors *Depression* and *Psychosis* were utilized as statistical tools for subtraction to isolate the genetic effects specific to *Mania*. The observed variables were bipolar disorder, schizophrenia, and MDD. The factor *Mania* was set to be loaded by bipolar disorder only, while the factor *Psychosis*, and the factor *Depression* by bipolar disorder and MDD, with factor loading coefficients allowed to be freely estimated. Following the original method by Demange et al. (20), the subtracting latent variables and *Mania* had their variances fixed to 1.00. In contrast to the model by Demange et al. (20) and to improve model accuracy due to shared genetic covariance between the underlying constructs of schizophrenia and MDD, a covariance path was added between the residuals of *Depression* and *Psychosis*. This was possible because our extended model included an additional degree of freedom, thanks to the extra indicator variable in our “double subtraction” model. Covariances among other residuals of the observed variables and between residuals of the latent variables were fixed to zero.

**Figure 1.**
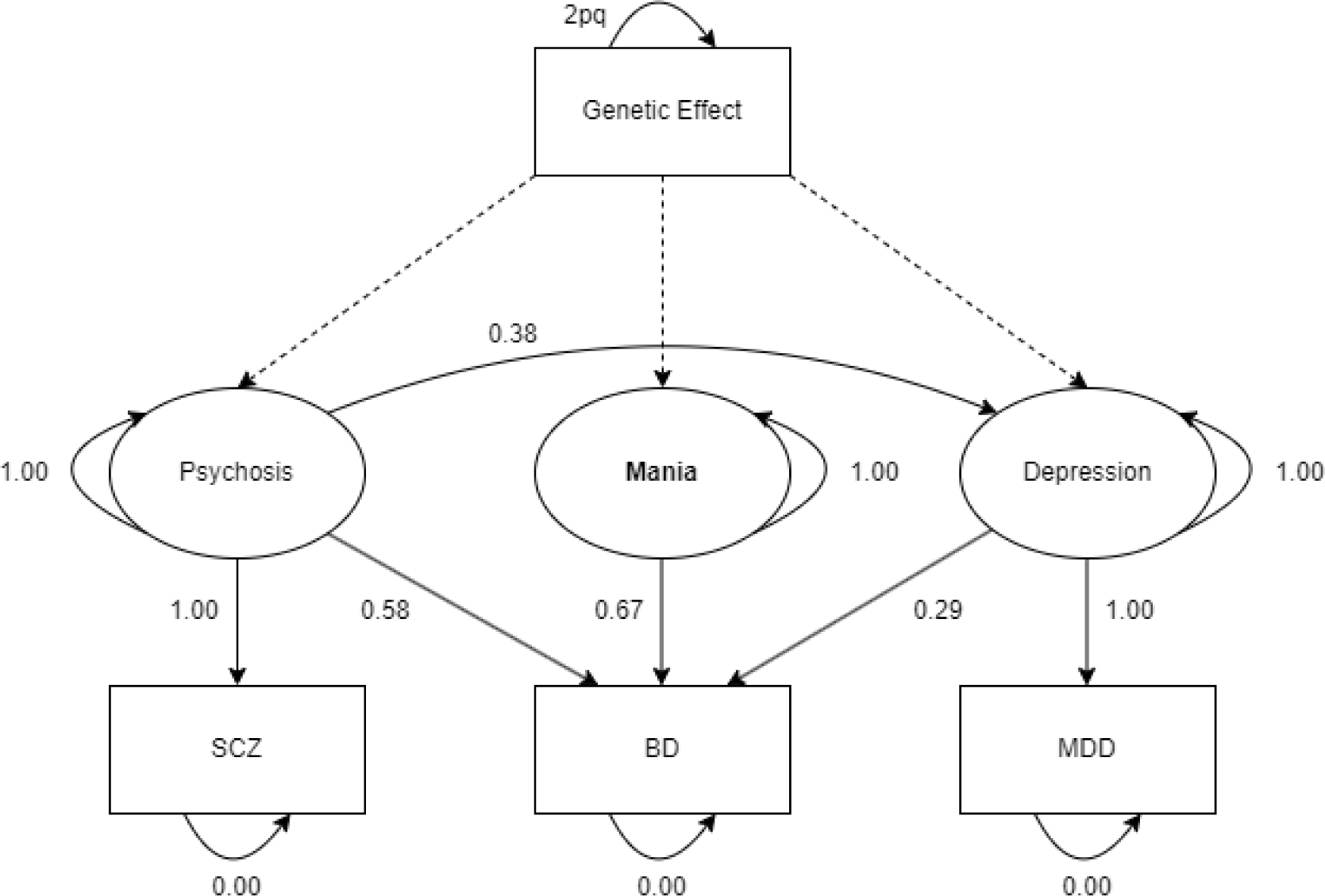
SEM path diagram. All SEM pathways are highly significant (p<0.001). Rectangles represent manifest variables while ellipses represent latent variables. Legend for traits: SCZ (schizophrenia), BD (bipolar disorder), and MDD (major depressive disorder). Dotted lines depict regression of genetic effects from SNPs to latent variables.

### GWAS of Latent Factors

Each SNP in the genome was regressed on *Mania* and on the subtracting latent variables, with the summary statistics harmonized with the 1000 Genomes reference set. This approach enabled three regression pathways for each SNP towards bipolar disorder: two mediated by *Psychosi*s and *Depression*, and one mediated by *Mania*. A graph of the model displaying factor loadings was also generated (Figure 1). Manhattan and QQ plots were generated for the *Mania* factor (Figure 2, Supplementary Figure 1 respectively).

**Figure 2.**
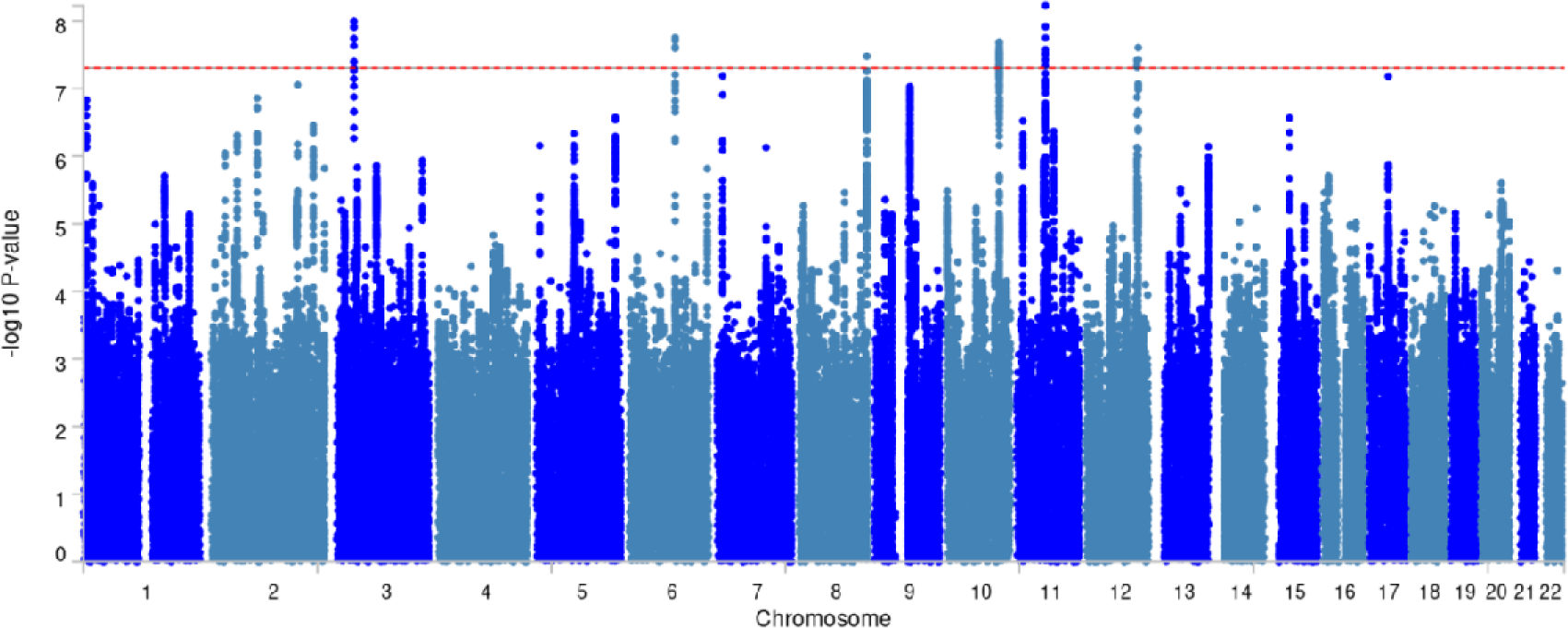
Manhattan plot of mania: genetic significance in synthetic GWAS analysis. The red line represents the threshold for genomic significance (5×10^-8^).

### Genetic Correlations

Genetic correlations of the *Mania* factor with psychiatric, substance abuse, social, somatic and neurological traits of interest (17,21,23,30–53) were explored using LD score regression (26) as implemented in GSEM. The correlations with mania were compared to those with bipolar disorder to determine if significant differences exist. To this aim, statistical tests for differences in genetic covariance and its variance were applied, accounting for the contributing number of genetic variants in the analysis (https://github.com/johanzvrskovec/shru). The test results were corrected for multiple comparisons using False Discovery Rate (FDR). Population and sample prevalence estimates for the studied trait were extracted from the original publication associated with the GWAS summary statistics.

### GWAS Locus Annotation

Functional Mapping and Annotation of Genome-Wide Association Studies, or FUMA (54) was utilized to perform LD clumping and detect lead SNPs and associated genes from the synthetic GWAS data on mania produced through GSEM (default settings were employed: estimation method = “DWLS”, tolerance = 1e-40, genomic control = “standard”).

Significantly associated variants (p□<□5×10^−8^) in LD clumps (r^2^ ≥ 0.6), were considered as lead SNPs. A second clumping step to single out independent significant lead SNPs was subsequently performed, selecting the stronger associated lead SNP in LD clumps of weaker correlation (r^2^≥0.1) within a maximum distance of 250 kb. Independent genome-wide significant loci defined in this way were then mapped to variant annotations in the NHGRI-EBI GWAS catalog.

## Results

### Double-GWAS-by-subtraction SEM Model

Significant factor loadings (Figure 1, Supplementary Table 1) on bipolar disorder for *Mania* (standardized coefficient: 0.67, p<0.001), *Psychosis* (0.58, p<0.001), and *Depression* (0.29, p<0.001), as well as a significant covariance path between *Psychosis* and *Depression* (0.38, p<0.001) were fitted in our investigated model. As the model is fully saturated, fit statistics are not available.

Heritability analysis for the mania phenotype yielded a mean □² of 1.152 across SNPs. The LDSC intercept was 0.91 (SE 0.006), and the LDSC SNP heritability (h²_SNP_) was 0.182 (SE 0.01).

### GWAS Locus Annotation

After FUMA analysis, 7 LD clumped peaks with one lead SNP were identified on chromosomes 3, 6, 8, 10, 11, 12 and 12 (Table 1, Figure 2). Three of the loci (rs9834970, 3q12.1; rs6992333, 8q13.1; rs174592, 11q13.1) were previously associated with bipolar disorder (17). The rest of the identified loci appear to be novel, with no prior reported associations with bipolar disorder, not even in the most recent and largest pre-published GWAS on bipolar (55). The peaks included three intergenic loci, two intronic loci, one non-coding RNA intronic locus, and one exonic locus (rs6992333).

**Table 1.**
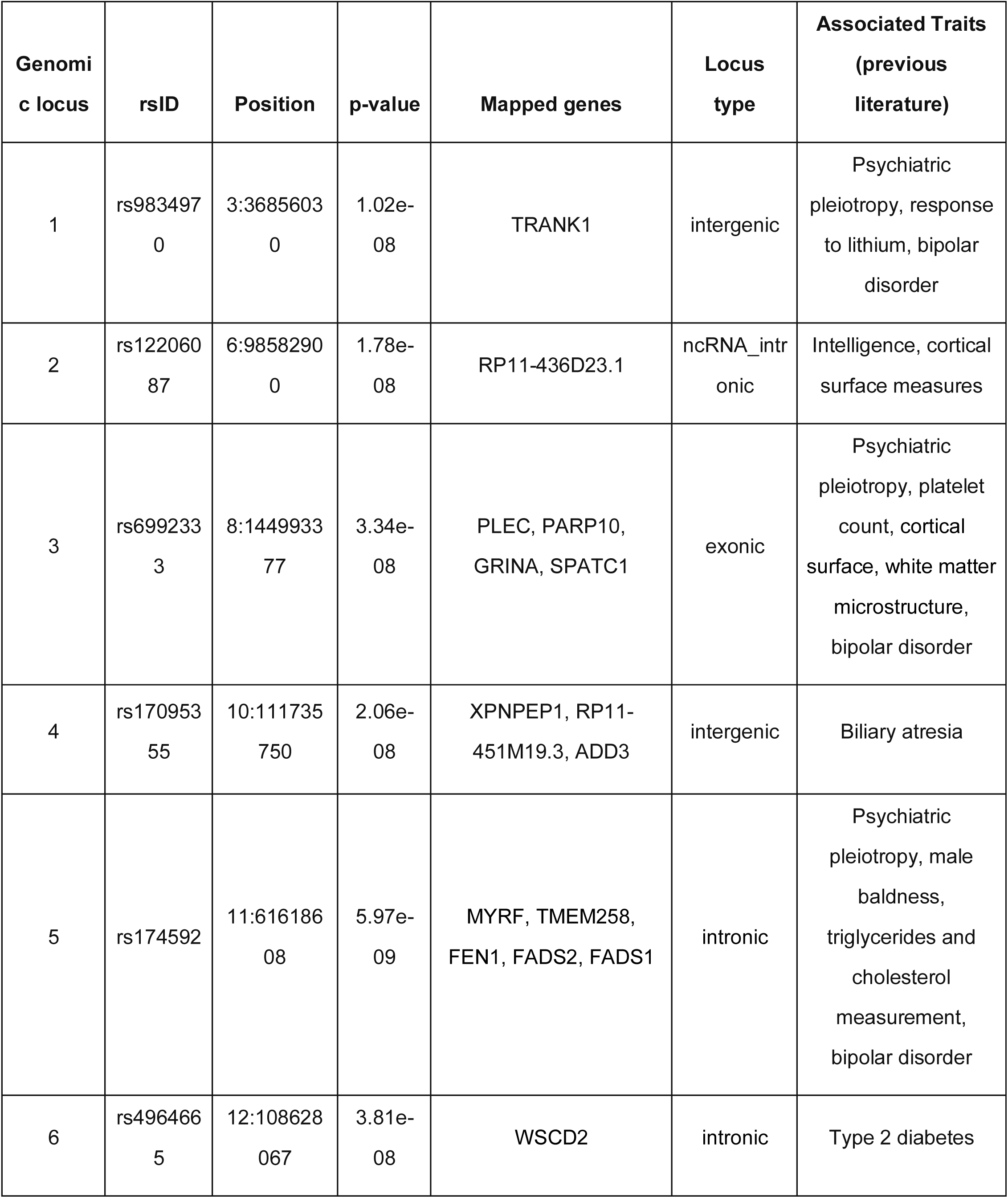
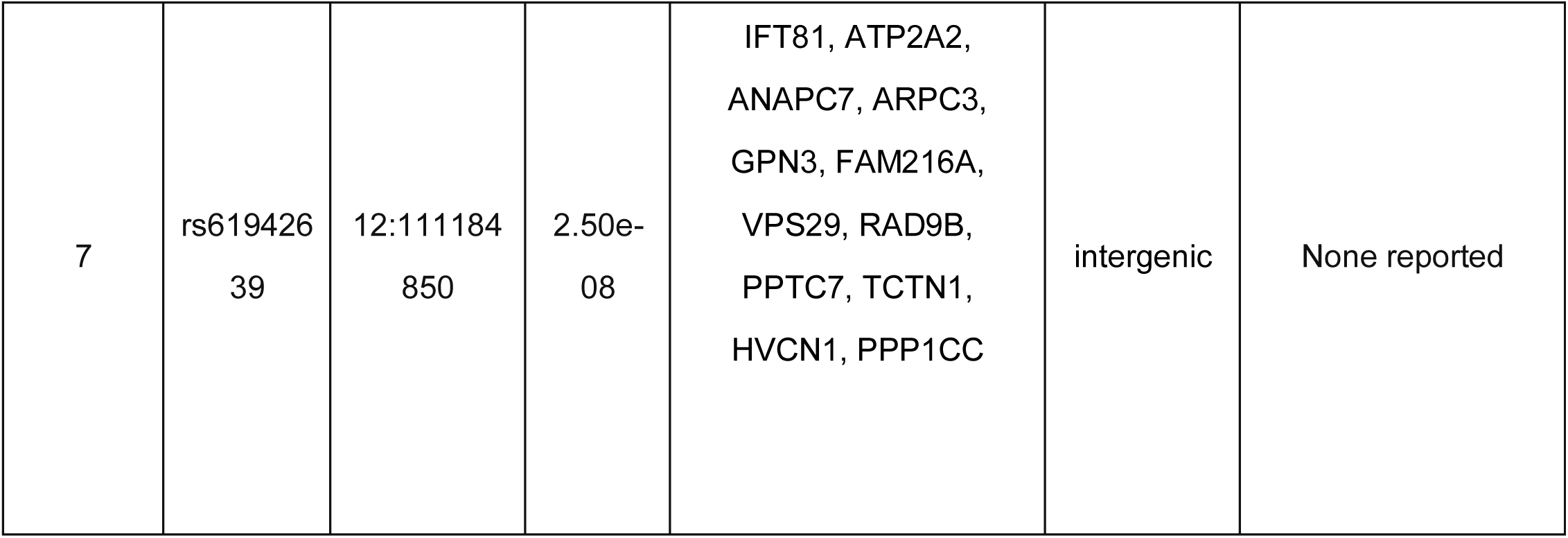
Genomic risk loci (bipolar disorder: BD)

### Genetic Correlations

Results of the genetic correlations with psychiatric, substance abuse and risky behaviors, social, somatic and neurological traits are shown in Figure 3 and Supplementary Table 2. The p-values presented in this analysis represent the statistical significance of the differences between the genetic correlation values (r_g_) for mania and bipolar disorder.

**Figure 3.**
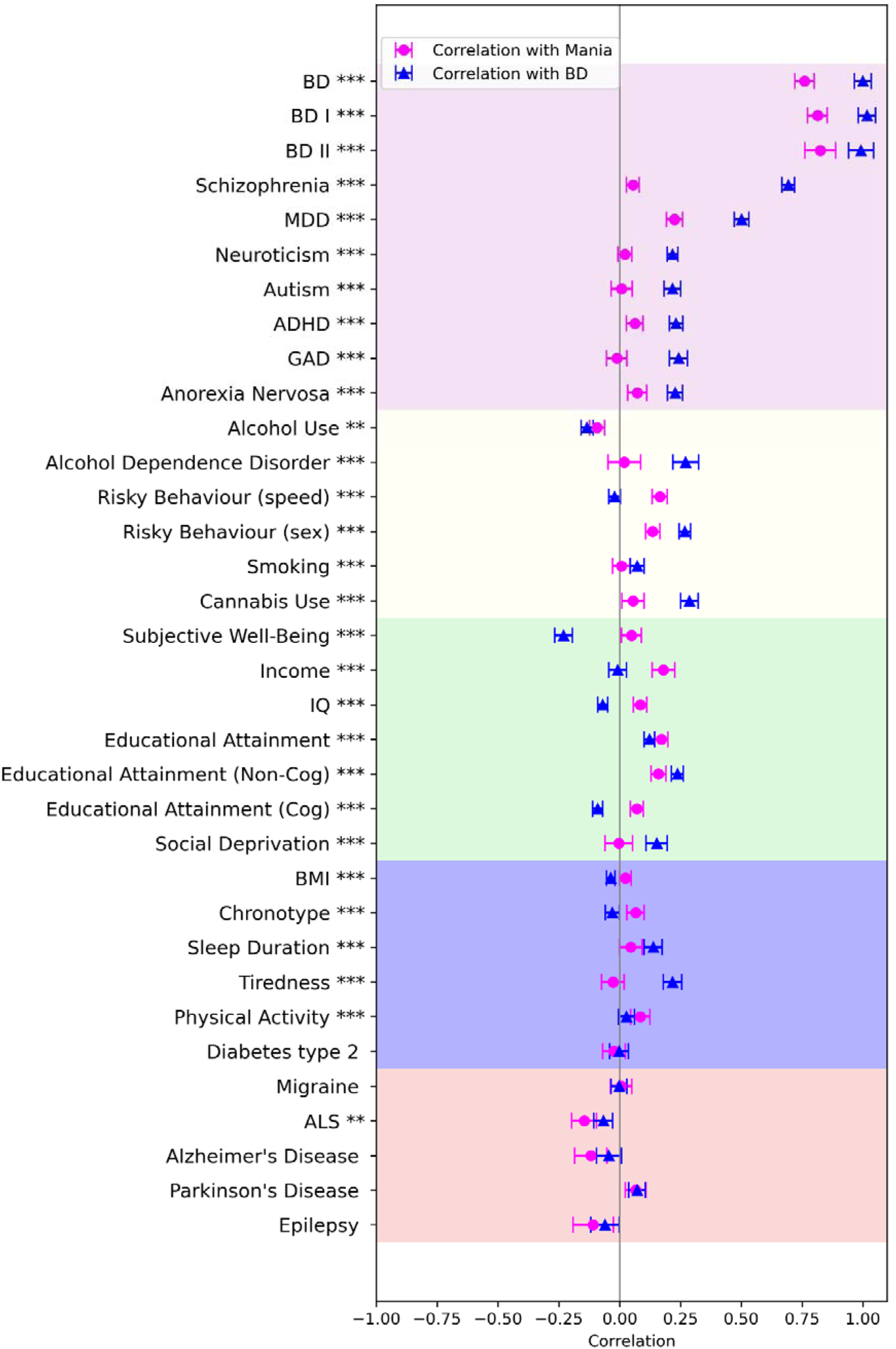
Genetic correlations of mania and bipolar disorder with psychiatric, substance abuse and risky behaviors, social, somatic, and neurological traits. Asterisks indicate significance levels of the difference between mania and bipolar disorder: * p<0.05, ** p<0.01, *** p<0.001.

Schizophrenia and MDD exhibited significantly lower (p<0.001) genetic correlations with mania (r_g_=0.05 for schizophrenia, r_g_=0.23 for MDD) than with bipolar disorder (r_g_=0.69 for schizophrenia, r_g_=0.50 for MDD), consistently with the model’s intent. Mania retained a high genetic correlation with bipolar disorder (r_g_=0.76). Genetic correlations of mania with other psychiatric conditions, such as autism (r_g_=0.01), ADHD (r_g_=0.06), neuroticism (r_g_=0.02), anorexia nervosa (r_g_=0.07), and generalized anxiety disorder (GAD, r_g_=0.01), were consistently lower compared to their correlations with bipolar disorder (r_g_=0.22 for autism, r_g_=0.23 for ADHD, r_g_=0.22 for neuroticism, r_g_=0.23 for anorexia nervosa, r_g_=0.24 for GAD). All differences in correlations were statistically significant (p<0.001).

Significant differences were also observed in correlations with substance abuse and impulsivity traits. Mania showed lower associations with substance abuse (cannabis use, r_g_=0.06; alcohol dependence, r_g_=0.02; smoking, r_g_=0.01) and risky sexual behavior, but an increased correlation with speeding, compared to bipolar disorder (differences in correlations with p<0.001).

Mania exhibited higher (p<0.001) genetic correlations than bipolar disorder with factors such as income (mania: r_g_=0.18, bipolar: r_g_=-0.01), physical activity (mania: r_g_=0.09, bipolar: r_g_=0.03), and subjective well-being (mania: r_g_=0.05, bipolar: r_g_=-0.23). Conversely, while bipolar disorder had a modest genetic correlation with social deprivation (bipolar: r_g_=0.15), this was reduced (p<0.001) in mania (mania: r_g_ < 0.001). Mania showed an increased (p<0.001) genetic correlation with the cognitive component of educational attainment (mania: r_g_=0.07, bipolar: r_g_=-0.09), but a significantly reduced (p<0.001) correlation with the non-cognitive component (mania: r_g_=0.16, bipolar: r_g_=0.24).

Somatic traits generally exhibited lower genetic correlations with both bipolar disorder and mania in terms of absolute values. However, statistically significant differences were still observed in specific traits such as BMI (p<0.001), where mania showed a slightly positive correlation (r_g_=0.02) compared to a negative correlation for bipolar disorder (r_g_=-0.04). For “morning” chronotype, which describes individuals who are more active and alert in the early hours of the day, mania had a positive correlation (r_g_=0.07), whereas bipolar disorder had a negative correlation (r_g_=-0.03). High sleep duration showed a positive correlation for both, but was higher for bipolar disorder (mania: r_g_=0.05, bipolar: r_g_=0.14). Tiredness was negatively correlated with mania (r_g_=-0.03) but positively correlated with bipolar disorder (r_g_=0.22). Physical activity showed a higher (p<0.001) positive correlation with mania (r_g_=0.09) compared to bipolar disorder (r_g_=0.03). The difference in correlation with type 2 diabetes was not statistically significant (p=0.175).

Neurological traits did not exhibit any statistically significant differences in correlation between mania and bipolar disorder, except for ALS (p<0.001) and Alzheimer’s disease (p=0.003). For ALS, mania showed a stronger negative correlation (r_g_=-0.15) compared to bipolar disorder (r_g_=-0.07,). Similarly, for Alzheimer’s disease, mania exhibited a more pronounced negative correlation (r_g_=-0.12) than bipolar disorder (r_g_=-0.04).

## Discussion

The present study leveraged a novel Genomic Structural Equation Modeling (GSEM) GWAS-by-subtraction approach to dissect the genetic components of bipolar disorder, specifically focusing on a latent factor representing mania. Our SEM model results underscore the central role of mania in the genetic construct of bipolar disorder, as indicated by the high standardized coefficient (0.67). Psychosis emerged as the second most significant component (coefficient: 0.58). Further, by utilizing this ‘double subtraction’ method, we were able to conduct a GWAS of mania and to calculate its genetic correlations. Our findings offer valuable insights into the genetic architecture of bipolar disorder and the unique genetic profile of mania.

The correlation analyses demonstrated distinct genetic correlation profiles between mania and bipolar disorder across various psychiatric, substance use, social, somatic, and neurological traits. These findings underscore the specific genetic components captured by the *Mania* factor, which appear to be significantly distinct from bipolar disorder (Supplementary Table 2). The attenuated genetic correlations between mania and psychiatric conditions align with our expectations of how the model’s structure would influence the latent *Mania* factor. Psychiatric conditions typically have high genetic correlations with each other. Therefore, by subtracting the effects of MDD and schizophrenia from the *Mania* factor, these correlations are reduced. This decrease suggests that the model effectively captures and adjusts for the shared genetic components among these conditions, supporting the latent structure imposed on the *Mania* factor.

The GWAS of *Mania* revealed seven genomic loci (Table 1). Several loci were previously already associated with multiple psychiatric conditions and bipolar disorder, including rs9834970 on chromosome 3q12.1 which has also been associated with lithium response (56). Locus 3 (rs6992333, 8q13.1) is linked not only to bipolar disorder but also to cortical surface, white matter microstructure and platelet counts (17,52,57,58). Locus 5 (rs174592, 11q13.1) has been connected with bipolar disorder, general psychiatric risk, cholesterol, triglyceride measurements, and male baldness (17,59–62). Among the loci not previously associated with bipolar disorder, rs4964665 (12q14.1) has been associated with type 2 diabetes (63), rs17095355 (10q21.1) to biliary atresia (64), and rs12206087 (6q14.3) to intelligence and cortical surface measures (57,65).

Clinical practice and the literature suggest that mania can be associated with bursts of high productivity in the workplace or academic settings for some individuals (66–68). While bipolar disorder as a whole is associated with lower performance in these areas, this effect is often attributed primarily to depressive symptoms (69–72). This may explain why the higher genetic correlations with income (mania: r_g_=0.16, bipolar: (r_g_=-0.01), IQ (mania: r_g_=0.16, bipolar: (r_g_=-0.07), and educational attainment (mania: r_g_=0.17, bipolar: (r_g_=0.12) with *Mania*. Interestingly, the non-cognitive part of educational attainment, which is largely theorized as being composed of traits like socio-economic status (SES), perseverance and motivation (43), showed reduced correlation with *Mania* (mania: r_g_=0.16, bipolar: r_g_=0.24). The removal of MDD and schizophrenia from the analysis might have eliminated some basic underlying vulnerability components (73) from the *Mania* factor, as well as part of the variance that would have contributed to residuals if allowed. In fact, some lines of research suggest that there might be some level of (sub)clinical impairment associated with schizophrenia and MDD (74,75). Additionally, evidence from mendelian randomization (MR) studies suggest that the relationship between low IQ and bipolar disorder might be non causal, and potentially mediated by the genetic component of schizophrenia (76).

*Mania* demonstrated higher correlations with traits such as physical activity, risky speeding behaviour, evening chronotype, and subjective well-being, and lower correlations with tiredness and sleep duration, compared to bipolar disorder. These differences in correlations align closely with mania’s symptomatology (3). The elevated correlations with traits like increased physical activity (mania: r_g_=0.09, bipolar disorder: r_g_=0.03) and risky speeding behaviour (mania: r_g_=0.16, bipolar disorder: r_g_=-0.02) and the negative correlation with tiredness (mania: r_g_=-0.03, bipolar disorder: r_g_=0.22) reflect the heightened energy levels and impulsivity typical of manic episodes (77). Similarly, the association with evening chronotype (mania: r_g_=0.07, bipolar disorder: r_g_=-0.03) and reduced sleep duration (mania: r_g_=0.05, bipolar disorder: r_g_=0.14) reflects the altered sleep patterns characteristic of mania (78,79).

Mania exhibited reduced genetic correlations with substance abuse disorders compared to BD, as seen with cannabis use (rg=0.06 for mania vs. 0.29 for BD), tobacco smoking (rg=0.01 for mania vs. 0.07 for BD), and alcohol dependence (rg=0.02 for mania vs. 0.27 for BD). Interestingly, mania showed increased correlations with traits associated with impulsivity and risk-taking, such as risky speeding behavior (rg=0.16 for mania vs. −0.02 for BD).

The relationship between mania and substance use appears more nuanced when examining alcohol consumption. While mania had a weaker correlation with alcohol dependence (a measure of problematic use), it showed a less negative correlation with general alcohol use (rg=-0.09 for mania vs. −0.13 for BD). This suggests that the genetic factors underlying mania might be less associated with addictive patterns of substance use, but rather potentially with impulsive behaviour more broadly. Interestingly, mania exhibited a reduced correlation with “risky sexual behaviour” (mania: r_g_=0.14, bipolar disorder: r_g_=0.27), defined as the self-reported number of sexual partners standardised for self-reported age and gender (37), contrasting with the hypersexual behaviour typically seen in bipolar patients (80). The number of sexual partners is influenced by various factors beyond impulsivity, such as social norms, personal relationships, and individual preferences (81), thereby partially explaining the diminished correlation with mania. An alternative explanation for the reduced correlation between mania and risky sexual behaviour involves the genetic architecture of mixed states in bipolar disorder. Mixed states, combining manic and depressive features, are associated with increased risky sexual behaviours (80). By removing the genetic component associated with depression from bipolar disorder, we may inadvertently remove factors contributing to mixed states. This could exclude genetic variants relevant to risky sexual behaviour in bipolar disorder, resulting in a lower genetic correlation between mania alone and risky sexual behaviour compared to bipolar disorder as a whole.

Finally, it must be noted that our mania factor is less genetically associated with lower SES as compared to bipolar disorder. The relationship between number of sexual partners and SES is complex and non-linear (82), with higher partner counts observed in both the lowest and highest SES quintiles. The UK Biobank, the primary data source for the summary statistics that we employed, underrepresents the lower SES quintiles (83). Consequently, the reduced genetic correlation between mania and lifetime sexual partner count may be an artifact of removing the genetic components associated with depression and schizophrenia, which are more strongly linked to lower SES. This removal could disproportionately affect the representation of lower SES quintiles in our analysis, potentially influencing the observed genetic correlations.

*Mania* mostly followed the correlation values of bipolar disorder regarding neurological traits, except in the cases of Alzheimer’s disease and ALS, where it exhibited significantly stronger negative genetic correlations (mania: r_g_=-0.12, bipolar disorder: r_g_=-0.04 for Alzheimer’s disease; mania: r_g_=-0.15, bipolar disorder: r_g_=-0.07 for ALS). Schizophrenia and MDD have been associated with neurotoxicity, and there is evidence supporting the notion that the respective treatments might have a positive impact on neurogenesis on both animal models and patients (84–87). Furthermore, lithium, a common treatment for bipolar disorder, has been proposed as a therapeutic agent for both Alzheimer’s disease (88) and ALS (89), although its efficacy in ALS is debated (90). Lithium’s effect in this context is mostly thought to be mediated by its neuroprotective properties (89). Thus, the removal of schizophrenia and MDD from the genetic component of bipolar might thus induce the increased negative genetic correlation between mania and Alzheimer’s disease, ALS and epilepsy (mania: r_g_=-0.11, bipolar disorder: r_g_=-0.06), all of which involve neurotoxicity as a pathogenetic mechanism (91–93).

While the present study provides valuable insights into the genetic structure of mania, several limitations must be acknowledged. Firstly, the model specification and assumptions imposed certain constraints that may not fully capture the intricate genetic architecture and complex relationships between the underlying constructs of mania, psychosis, and depression; as an example, due to missing degrees of freedom, the only covariance path that could be allowed between these variables was between *Depression* and *Psychosis*. This imposition, while grounded in theoretical considerations (94), could have failed to capture all the nuances of the relations between the variables. Secondly, the use of GWAS summary statistics from European ancestry cohorts potentially limits the generalizability of the findings to other populations (95); thus, the current results may not accurately reflect the genetic structure of mania in non-European ethnicities. Additionally, the average Chi-square values varied across conditions, with the MDD sample showing a significantly lower mean Chi-square (1.2006) compared to schizophrenia (2.1224) and bipolar disorder (1.5886), which may reflect differences in sample sizes and the detectability of genetic signals. Since the model is fully saturated, fit statistics are not available, making it difficult to assess the model’s quality. Finally, since the GSEM package does not support GWAS analysis for the X chromosome, data from the X chromosome were excluded from the analysis.

In conclusion, we demonstrate the use of a double GWAS-by-subtraction model allows the isolation of a *Mania* latent factor. The GWAS of *Mania* identified 7 significant genomic loci, four of which were not previously associated with bipolar disorder. Genetic correlation analysis revealed that *Mania* has a unique correlation profile compared to bipolar disorder. As expected, *Mania* exhibited lower genetic correlations with most psychiatric conditions due to removal of effects for both schizophrenia and bipolar disorder. However, it showed distinct correlations with traits such as physical activity, subjective well-being, sleep duration, chronotype and income, reflecting the behavioural and psychopathological manifestations of mania. This study demonstrates the effectiveness of GSEM in dissecting the complex genetic structure of bipolar disorder, offering a precision-medicine pathway to more precise genetic risk prediction and targeted treatments, aimed at specific endophenotypes.

## Data Availability

No data was produced or gathered for the current study. The code used in the study can be shared upon reasonable request.

## Declarations

### Availability of data and code

No data was produced or gathered for the current study. The code used in the study can be found at the following GitHub repository: https://github.com/tnggroup/GSEM-DoubleSub. The study was pre registered at https://osf.io/7ka93.

### Authors Contribution

## Abbreviations

MDD: major depressive disorder
SEM: structural equation modeling
GSEM: genomic structural equation modeling
GWAS: genome-wide association study
SNP: single nucleotide polymorphism
BD: bipolar disorder
MAF: minor allele frequency
LDSC: linkage disequilibrium score regression
FDR: false discovery rate
FUMA: functional mapping and annotation
ALS: amyotrophic lateral sclerosis
ADHD: attention-deficit/hyperactivity disorder
IQ: intelligence quotient
GAD: generalized anxiety disorder

## Supplementary Figure Legends

**Supplementary Figure 1.**
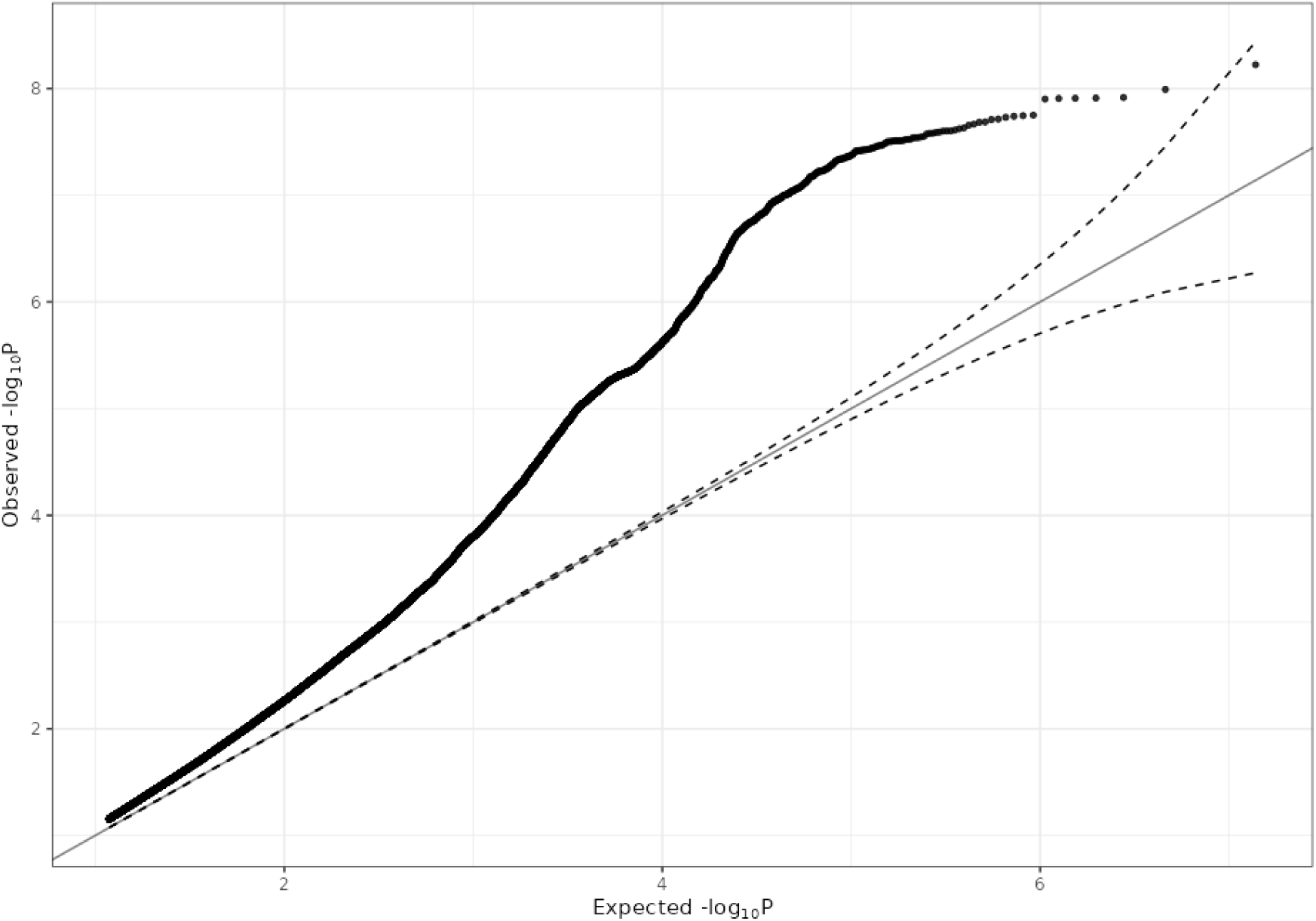
QQ plot of the Mania factor: comparison of p-values from observed and theoretical distributions (λ=1.115).

**Supplementary Table 1.**
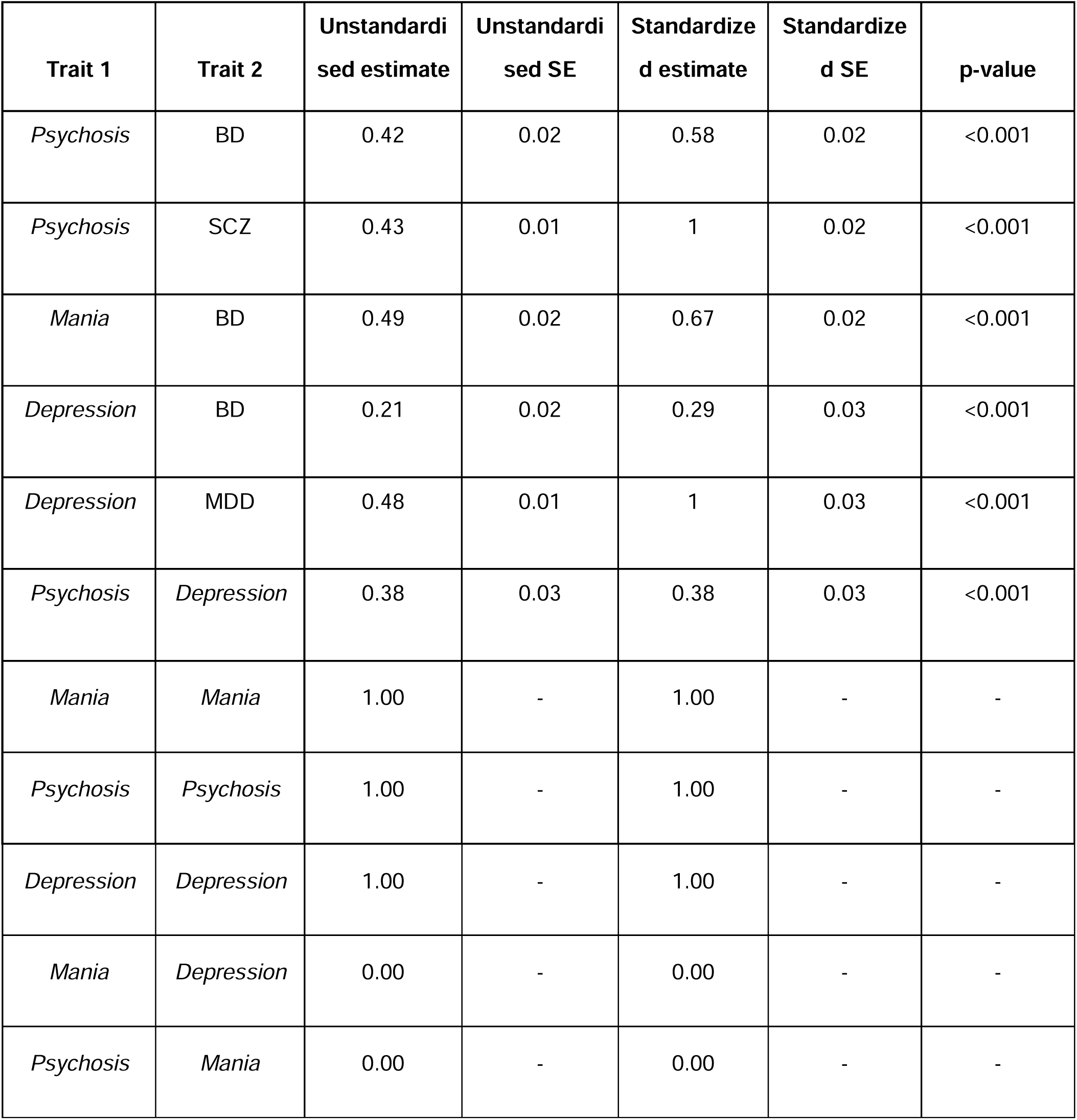
SEM estimates (schizophrenia: SCZ, bipolar disorder: BD, major depressive disorder: MDD, standard error: SE)

**Supplementary Table 2.**
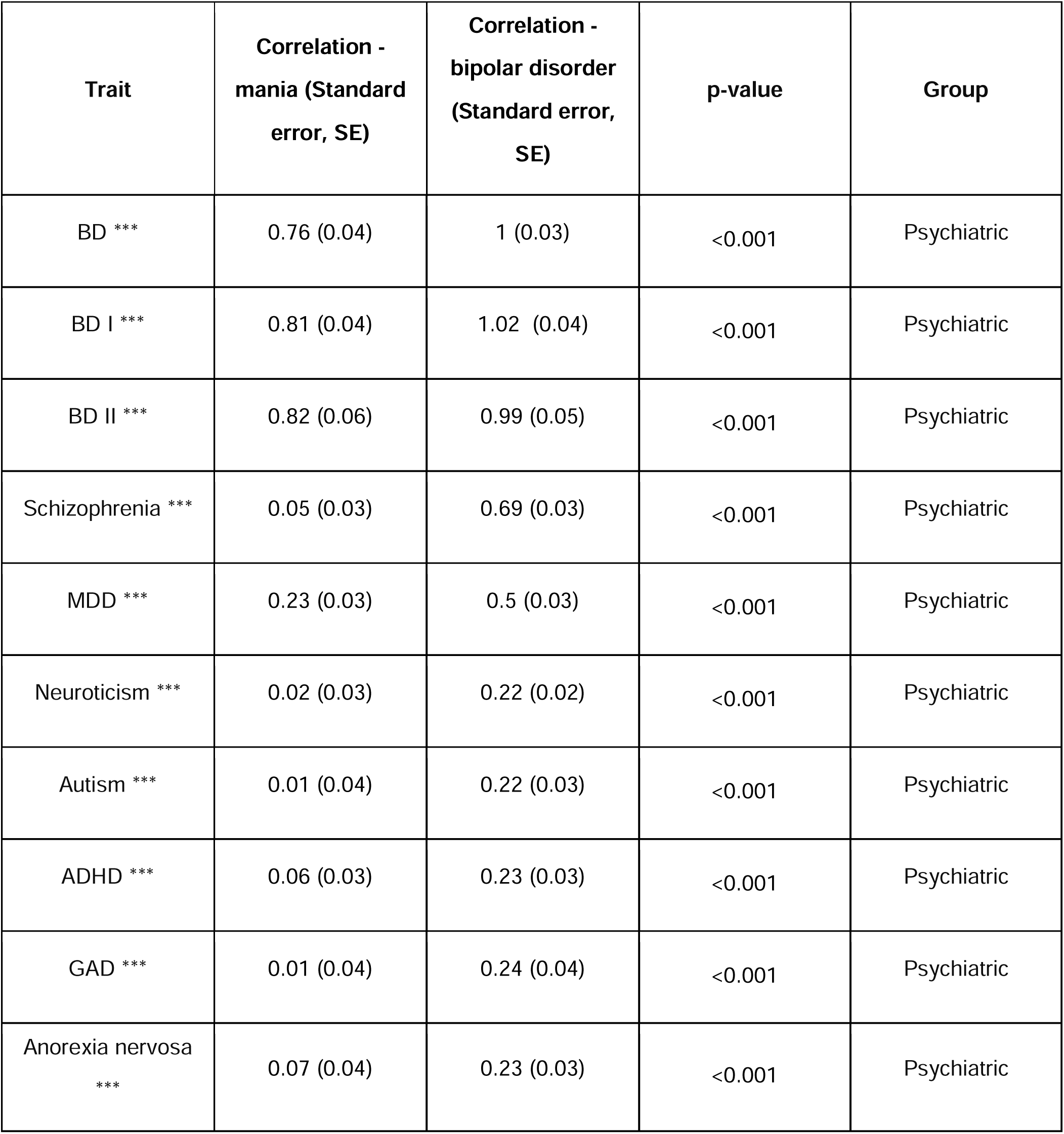
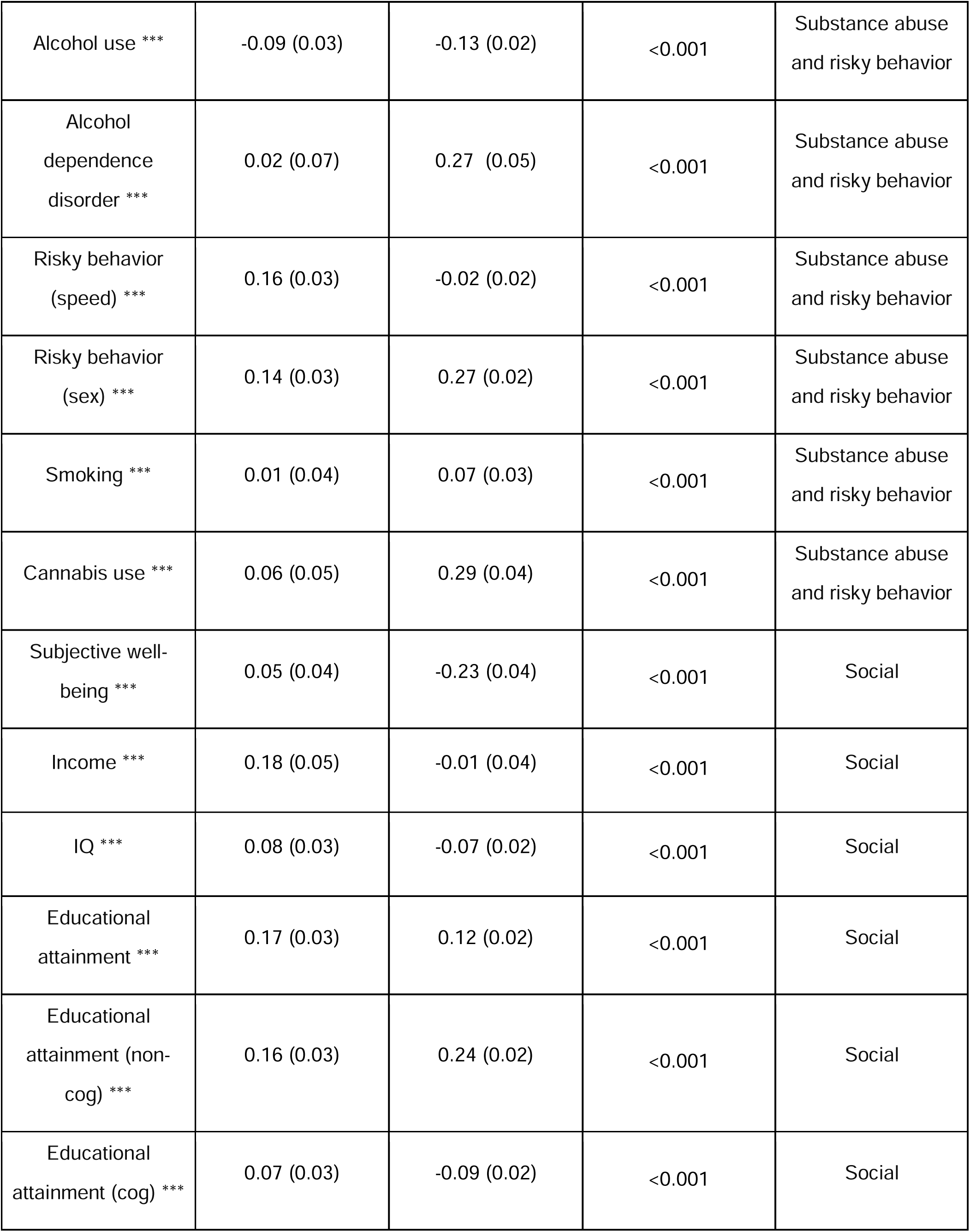
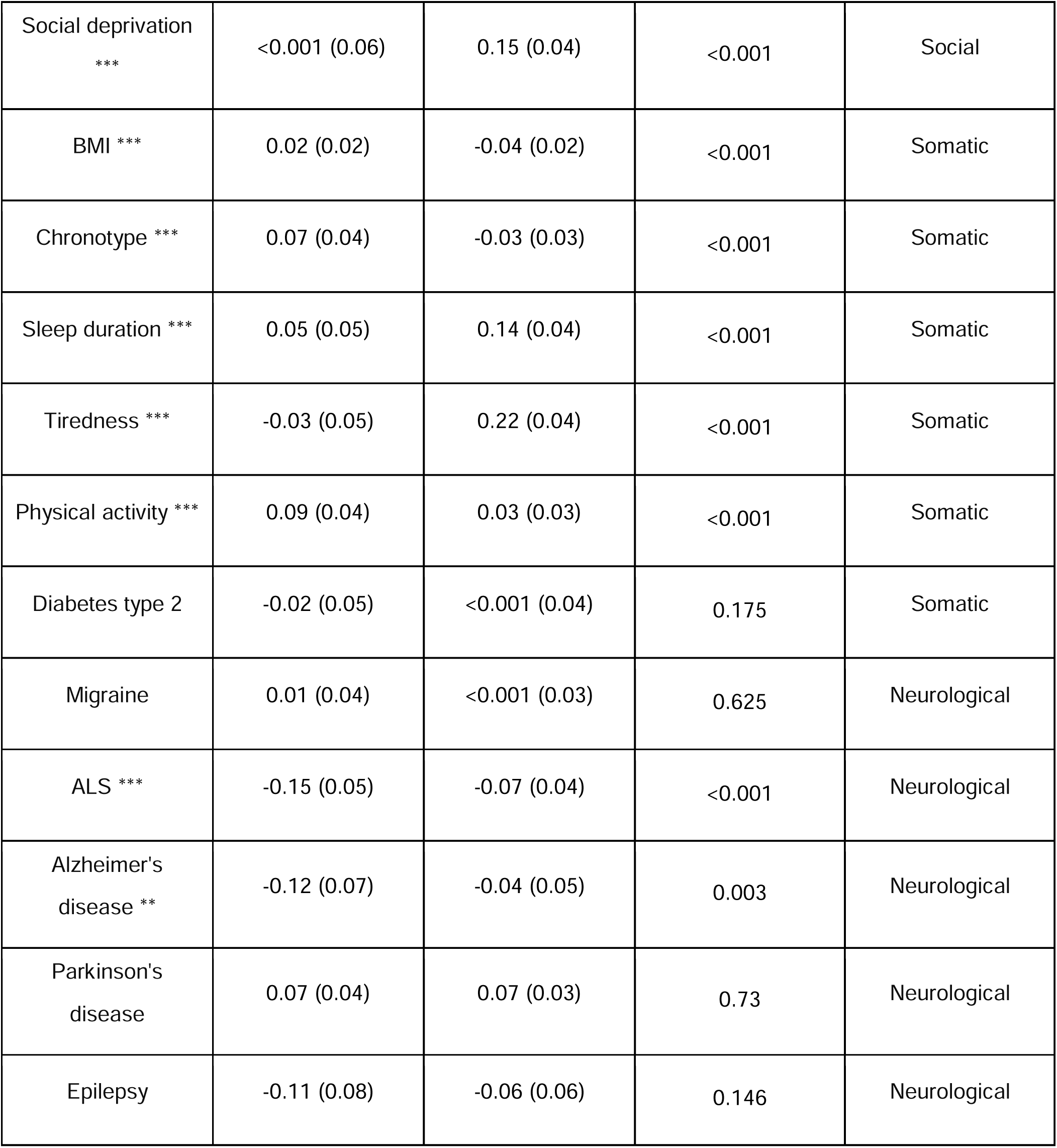
Genetic correlations of mania and bipolar disorder with psychiatric, substance abuse, social, somatic, and neurological traits (* p<0.05, ** p<0.01, *** p<0.001, FDR adjusted). Bipolar disorder type I: BD I, bipolar disorder type II: BD II, generalized anxiety disorder: GAD, intelligence quotient: IQ, body mass index: BMI, amyotrophic lateral sclerosis: ALS.

