## Supplementary Table 2 for "Isolating the genetic component of mania in bipolar disorder"

### ***Supplementary Table 2 -*** *Genetic correlations of mania and bipolar disorder with psychiatric, substance abuse, social, somatic, and neurological traits (* p<0.05, ** p<0.01, *** p<0.001, FDR adjusted). Bipolar disorder type I: BD I, bipolar disorder type II: BD II, generalized anxiety disorder: GAD, intelligence quotient: IQ, body mass index: BMI, amyotrophic lateral sclerosis: ALS.*

| **Trait** | **Correlation - mania (Standard error, SE)** | **Correlation - bipolar disorder (Standard error, SE)** | **p-value** | **Group** |
| --- | --- | --- | --- | --- |
| BD *** | 0.76 (0.04) | 1 (0.03) | <0.001 | Psychiatric |
| BD I *** | 0.81 (0.04) | 1.02 (0.04) | <0.001 | Psychiatric |
| BD II *** | 0.82 (0.06) | 0.99 (0.05) | <0.001 | Psychiatric |
| Schizophrenia *** | 0.05 (0.03) | 0.69 (0.03) | <0.001 | Psychiatric |
| MDD *** | 0.23 (0.03) | 0.5 (0.03) | <0.001 | Psychiatric |
| Neuroticism *** | 0.02 (0.03) | 0.22 (0.02) | <0.001 | Psychiatric |
| Autism *** | 0.01 (0.04) | 0.22 (0.03) | <0.001 | Psychiatric |
| ADHD *** | 0.06 (0.03) | 0.23 (0.03) | <0.001 | Psychiatric |
| GAD *** | 0.01 (0.04) | 0.24 (0.04) | <0.001 | Psychiatric |
| Anorexia nervosa *** | 0.07 (0.04) | 0.23 (0.03) | <0.001 | Psychiatric |
| Alcohol use *** | -0.09 (0.03) | -0.13 (0.02) | <0.001 | Substance abuse and risky behavior |
| Alcohol dependence disorder *** | 0.02 (0.07) | 0.27 (0.05) | <0.001 | Substance abuse and risky behavior |
| Risky behavior (speed) *** | 0.16 (0.03) | -0.02 (0.02) | <0.001 | Substance abuse and risky behavior |
| Risky behavior (sex) *** | 0.14 (0.03) | 0.27 (0.02) | <0.001 | Substance abuse and risky behavior |
| Smoking *** | 0.01 (0.04) | 0.07 (0.03) | <0.001 | Substance abuse and risky behavior |
| Cannabis use *** | 0.06 (0.05) | 0.29 (0.04) | <0.001 | Substance abuse and risky behavior |
| Subjective well-being *** | 0.05 (0.04) | -0.23 (0.04) | <0.001 | Social |
| Income *** | 0.18 (0.05) | -0.01 (0.04) | <0.001 | Social |
| IQ *** | 0.08 (0.03) | -0.07 (0.02) | <0.001 | Social |
| Educational attainment *** | 0.17 (0.03) | 0.12 (0.02) | <0.001 | Social |
| Educational attainment (non-cog) *** | 0.16 (0.03) | 0.24 (0.02) | <0.001 | Social |
| Educational attainment (cog) *** | 0.07 (0.03) | -0.09 (0.02) | <0.001 | Social |
| Social deprivation *** | <0.001 (0.06) | 0.15 (0.04) | <0.001 | Social |
| BMI *** | 0.02 (0.02) | -0.04 (0.02) | <0.001 | Somatic |
| Chronotype *** | 0.07 (0.04) | -0.03 (0.03) | <0.001 | Somatic |
| Sleep duration *** | 0.05 (0.05) | 0.14 (0.04) | <0.001 | Somatic |
| Tiredness *** | -0.03 (0.05) | 0.22 (0.04) | <0.001 | Somatic |
| Physical activity *** | 0.09 (0.04) | 0.03 (0.03) | <0.001 | Somatic |
| Diabetes type 2 | -0.02 (0.05) | <0.001 (0.04) | 0.175 | Somatic |
| Migraine | 0.01 (0.04) | <0.001 (0.03) | 0.625 | Neurological |
| ALS *** | -0.15 (0.05) | -0.07 (0.04) | <0.001 | Neurological |
| Alzheimer's disease ** | -0.12 (0.07) | -0.04 (0.05) | 0.003 | Neurological |
| Parkinson's disease | 0.07 (0.04) | 0.07 (0.03) | 0.73 | Neurological |
| Epilepsy | -0.11 (0.08) | -0.06 (0.06) | 0.146 | Neurological |
