## Supplementary Table 1 for "Isolating the genetic component of mania in bipolar disorder"

### ***Supplementary Table 1 -*** *SEM estimates (schizophrenia: SCZ, bipolar disorder: BD, major depressive disorder: MDD, standard error: SE)*

| **Trait 1** | **Trait 2** | **Unstandardised estimate** | **Unstandardised SE** | **Standardized estimate** | **Standardized SE** | **p-value** |
| --- | --- | --- | --- | --- | --- | --- |
| *Psychosis* | BD | 0.42 | 0.02 | 0.58 | 0.02 | <0.001 |
| *Psychosis* | SCZ | 0.43 | 0.01 | 1 | 0.02 | <0.001 |
| *Mania* | BD | 0.49 | 0.02 | 0.67 | 0.02 | <0.001 |
| *Depression* | BD | 0.21 | 0.02 | 0.29 | 0.03 | <0.001 |
| *Depression* | MDD | 0.48 | 0.01 | 1 | 0.03 | <0.001 |
| *Psychosis* | *Depression* | 0.38 | 0.03 | 0.38 | 0.03 | <0.001 |
| *Mania* | *Mania* | 1.00 | - | 1.00 | - | - |
| *Psychosis* | *Psychosis* | 1.00 | - | 1.00 | - | - |
| *Depression* | *Depression* | 1.00 | - | 1.00 | - | - |
| *Mania* | *Depression* | 0.00 | - | 0.00 | - | - |
| *Psychosis* | *Mania* | 0.00 | - | 0.00 | - | - |
